# Salivary profiles of 11-oxygenated androgens follow a diurnal rhythm in patients with congenital adrenal hyperplasia

**DOI:** 10.1101/2021.02.02.21249720

**Authors:** Hanna Nowotny, Matthias K. Auer, Christian Lottspeich, Heinrich Schmidt, Ilja Dubinski, Martin Bidlingmaier, Jo Adaway, James Hawley, Brian Keevil, Nicole Reisch

## Abstract

**Context:** Several studies have highlighted the importance of the 11-oxygenated 19-carbon (11oxC19) adrenal-derived steroids as potential biomarkers for monitoring patients with 21-hydroxylase deficiency (21OHD).

**Objective:** To analyze circadian rhythmicity of 11oxC19 steroids in saliva profiles and evaluate their relevance as potential monitoring parameters in 21OHD.

**Design, Setting, and Participants:** Cross-sectional single center study including 34 patients with classic 21OHD (men=14; women=20) and 32 BMI- and age-matched controls (men=15; women=17).

**Outcome Measures:** Salivary concentrations of the following steroids were analyzed by LC-MS/MS: 17-hydroxyprogesterone (17OHP), androstenedione (A4), testosterone (T), 11β-hydroxyandrostenedione (11OHA4) and 11-ketotestosterone (11KT).

**Results:** Similar to the previously described rhythmicity of 17OHP, 11OHA4 and 11KT concentrations followed a distinct diurnal rhythm in both patients and controls with highest concentrations in the early morning and declining throughout the day (**11-OHA4:** male patients Δ_mean_ = 79 %; male controls Δ_mean_ = 81%; female patients Δ_mean_ = 33 %; female controls Δ_mean_ = 91 %; **11KT:** male patients Δ_mean_ = 64 %; male controls Δ_mean_ = 60 %; female patients Δ_mean_ = 49 %; female controls Δ_mean_ = 81 %). Significant correlations between the area under the curve (AUC) for 17OHP and 11KT (r(p)_male_ = 0.741**; r(p)_female_ = 0.842****), and 11OHA4 (r(p)_male_ = 0.385^n.s.^; r(p)_female_ = 0.527*) were observed in patients but not in controls.

**Conclusions:** Adrenal 11oxC19 androgens are secreted following a diurnal pattern. This should be considered when evaluating their utility for monitoring treatment control.

## Introduction

Classic congenital adrenal hyperplasia (CAH) secondary to 21-hydroxylase deficiency (21OHD) is characterized by insufficient cortisol synthesis; and, in approximately two-thirds of patients, aldosterone production is also affected. Patients affected by CAH require life-long glucocorticoid (GC) and, where appropriate, mineralocorticoid replacement therapy. Cortisol deficiency results in adrenocorticotropic hormone (ACTH) over-secretion due to loss of negative feedback along the hypothalamic-pituitary-axis (1), consequently resulting in adrenal androgen excess (2). Accumulation of steroids upstream of the enzymatic blockade is a hallmark of the disease. In 21OHD, steroid precursor concentrations, especially 17-hydroxyporgesterone (17OHP), are substantially elevated underlining its pivotal role in the diagnostic work-up. Serum 17OHP is also the traditional indicator of adequate GC treatment in 21OHD. While in the general population, 17OHP is not a relevant source for adrenal androgen secretion, its excessive formation plays an essential role in adrenal hyperandrogenism in 21OHD (3-5).

Monitoring GC treatment to ensure optimal therapeutic effect and preventing hypo- or hyper-androgenism is essential and a challenging task of clinical care in 21OHD. In addition to the assessment of clinical parameters, the current guideline highlights the importance of regular and consistently timed hormone measurements relative to medication schedules and time of the day, due to their diurnal variance and dependency with regard to timing of GC replacement (6).

Disease monitoring is typically performed by morning blood tests before or after the intake of morning medication. Earlier studies have confirmed a strong correlation between plasma and salivary steroids in 21OHD supporting the idea of using this non-invasive monitoring strategy (7-10). Other approaches to estimate overall GC and androgen exposure include measurement of urine steroid metabolites, (serial) blood spots or steroid profiling in hair as potential long-term assessment of disease control (11-14).

With regard to biomarkers, most clinicians measure serum 17OHP, androstenedione (A4) and testosterone (T) concentrations and/or their metabolites in urine to assess disease control (15). Additionally, 21-deoxycortisol (21DF) has been described as a promising marker for monitoring 21OHD (16,17).

Recently, it has also been found that the 11-oxygenated 19-carbon (11oxC19) androgens are a clinically relevant androgenetic source and potential disease marker in 21OHD patients (18). The 11oxC19 androgens are derived from A4 which can be converted to 11β-hydroxyandrostenedione (11OHA4) by 11β-hydroxylase (CYP11B1) and further to 11-ketoandrostenedione (11KA4) and 11-ketotestosterone (11KT) through the action of 11β-hydroxysteroid dehydrogenase type 2 (11β-HSD2) (Figure 1) (19). In addition, 11oxC19 androgens also derive from 21DF (12). Hence, concentrations of 11oxC19 steroids are significantly elevated in patients with classic 21OHD compared to age-matched controls (18). The importance of 11KT and its derivate 11-ketodihydrotestosterone (11KDHT) is highlighted by the fact that both act at the androgen receptor with equal potency to their classic androgen counterparts, T and dihydrotestosterone (DHT) respectively.

**Figure 1.**
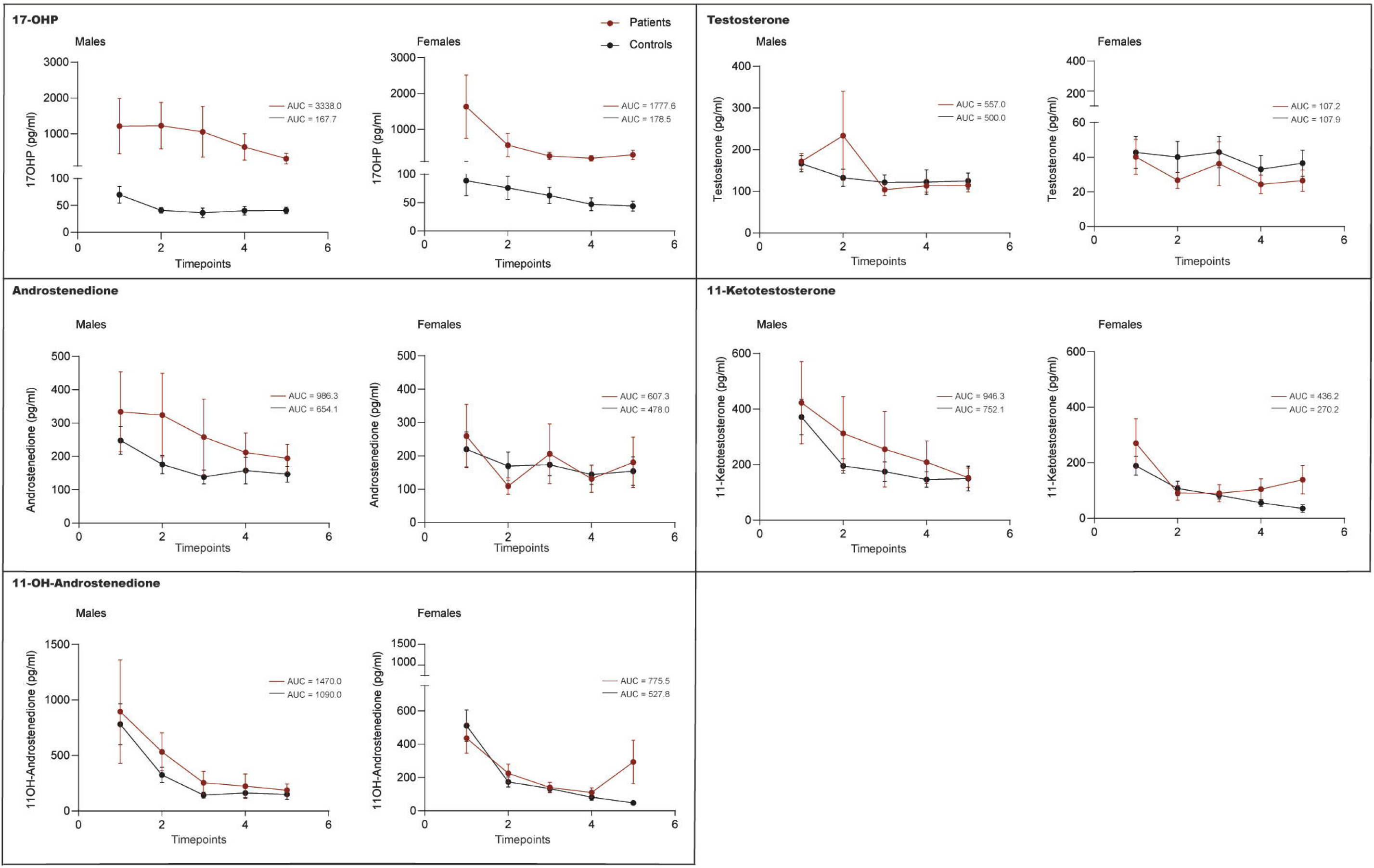
Pathway of 11oxC19 androgen synthesis. In patients with CAH due to 21OHD, 17OHP accumulates, which translates to A4 overproduction. A4 is converted to 11OHA4 by CYP11B1 and further to 11KT through the action of 11β-HSD2. 11oxC19 androgens also derive from 21DF.

As11oxC19 steroids are adrenal-specific androgens, they have been suggested as potentially novel biomarkers for adrenal androgen excess in 21OHD (17).

A recent publication showed a reliable correlation between morning plasma and salivary adrenal-specific androgens in 21OHD allowing a more comprehensive, non-invasive biomarker profiling in 21OHD (20).

Given the pivotal role of ACTH-mediated acute steroidogenic response, including upregulation of CYP11B1 and 11β-HSD2 (21) and the distinctive diurnal rhythm of ACTH (22) and 17OHP (23), a circadian secretion of 11oxC19 steroids seems obvious but has not been demonstrated so far. We hypothesized that salivary 11OHA4 and 11KT-levels would follow a diurnal profile with higher concentrations in the morning and lower concentrations in the evening in patients and controls. The current study investigated diurnal variation of adrenal-specific androgens via salivary steroid day profiles compared to controls as an effective non-invasive treatment monitoring tool.

## Subjects and Methods

### Subjects

Patients were recruited from the Endocrine Outpatient Clinic of the University Hospital Munich, Germany. All patients provided written informed consent to participate in our registry and biobank for adrenal insufficiency and differences of sex development (Bio AI/DSD, ethical approval no. 19-558). All patients had genetically and/or clinically confirmed classic CAH due to 21OHD. The control group was retrospectively selected from the control group of the German Cushing’s Registry (NeoExNET, ethical approval no. 152-10) after exclusion of Cushing’s syndrome based on a comparable anthropometric profile to the patient cohort (24). Exclusion criteria were defined as any condition or medication known to affect steroid metabolism as well as having worked in shift work during the last three months.

The study included 14 male and 20 female patients with CAH of whom 23 were suffering from salt wasting (SW) and 11 of the simply virilizing (SV) form of CAH. The control subjects were BMI- and age-matched (male, n = 15, female, n = 17).

Saliva samples were collected by carrying out saliva day profiles using saliva sampling via routinely used salivettes (Sarstedt, Nümbrecht (DE)) at five timepoints throughout the day. For control patients the following fixed timepoints were used: 08:00, 12:00, 16:00, 20:00 and 22:00 h. For CAH patients, timepoints of saliva sampling were adjusted to the intake of GC medication, so that the five samples were collected following the subsequent guidance: i) the first sample was collected upon awakening before intake of the morning GC dose, ii) the second sample at approximately 10:00 h, iii) the third sample at lunchtime, iv) the fourth sample in the afternoon at approximately 16:00 h and v) the last sample was collected before bedtime at approximately 22:00 h. All participants had previously been instructed on standardized use of salivettes.

One of the male and four of the female control patients were identified as outliers due to moderately elevated baseline 17OHP measurements and excluded from further analysis (Table 1). These were excluded from analysis and defined as outliers as lab parameters and original clinical parameters suspected carriers of 21OHD or non-classic 21OHD. The final data set included 14 male patients and 14 male controls as well as 20 female patients and 13 BMI- and age-matched controls. Data sets with three to four different timepoints of sample collection distributed throughout the whole day made up 20.0 to 42.9 % of the data sets analyzed. Incomplete data sets were mainly due to sporadic interference in data analysis or insufficient sample volume. The numbers and percentage of data points with values smaller than the lower limit of quantification (LLOQ) is listed per analyte. For values lower than the LLOQ, values were equated to the respective LLOQ (Table 1).

**Table 1.** Characteristics of data set.

| Variable | Men with CAH<br>(n=14) (%) | Control men<br>(n=15) (%) | Women with CAH<br>(n=20) (%) | Control women<br>(n=17) (%) |
| --- | --- | --- | --- | --- |
| Number of data points | 70 | 75 | 100 | 85 |
| Incomplete data sets | 6 (42.9) | 3 (20.0) | 6 (30.0) | 7 (41.2) |
| Outliers (elevated 17OHP) | NA | 1 (6.7) | NA | 4 (23.5) |
| 17OHP < LLOQ of 12.5 | NA | 8 (10.7) | 20 (20.0) | 14 (16.5) |
| 11KT < LLOQ of 6 | 8 (11.4) | NA | 24 (24.0) | 10 (11.8) |
| 11OHA4 < LLOQ of 40 | 11 (15.7) | 6 (8.0) | 27 (27.0) | 12 (14.1) |
| T < LLOQ of 5 | NA | NA | 30 (30.0) | 6 (7.1) |
| A4 < LLOQ of 10 | NA | NA | 17 (17.0) | 1 (1.2) |

Presentation of values as n (%). NA=not applicable.

**Table 2.** Assessment of therapy control of male and female patients with CAH.

| Variable | Men with CAH<br>(n=14) (%) | Women with CAH<br>(n=20) (%) |
| --- | --- | --- |
| AUC 17OHP | 50.8 – 433.1 $\frac{pg \times t(n)}{ml}$ | 25.0 – 450.5 $\frac{pg \times t(n)}{ml}$ |
| *Within reference range* | 6 (42.9) | 10 (50.0) |
| *Above reference range* | 8 (57.1) | 10 (50.0) |
| AUC A4 | 80.9 – 1518 $\frac{pg \times t(n)}{ml}$ | 57.7 – 1113.0 $\frac{pg \times t(n)}{ml}$ |
| *Within reference range* | 12 (85.7) | 17 (85.0) |
| *Above reference range* | 2 (14.3) | 3 (15.0) |
| AUC T | 112.9 – 1305.0 $\frac{pg \times t(n)}{ml}$ | 10.0 – 287.6 $\frac{pg \times t(n)}{ml}$ |
| *Within reference range* | 13 (92.9) | 18 (90.0) |
| *Above reference range* | 1 (7.1) | 2 (10.0) |
| AUC A4/T ratio | 0.72 – 2.76 | 1.83 – 12.45 |
| *Within reference range* | 12 (85.7) | 18 (90.0) |
| *Out of reference range* | 2 (14.3) | 2 (10.0) |

Presentation of values as n (%).

### Steroid hormone analysis

A validated LC-MS/MS assay was used for the simultaneous measurement of 17OHP, T, A4, 11OHA4 and 11KT (20,25). Standards and control solution containing a mixture of all analytes at the desired concentrations were prepared by dissolving the steroid in ultra-pure methanol and further dilution in a surrogate saliva matrix. The following analytes were used in the internal standard: [^13^C3]-(2,3,4)-17-alpha-hydroxyprogesterone, [^13^C3]-(2,3,4)-androstene-3,17-dione, [^13^C3]-(2,3,4)-testosterone, D7-2,2,4,6,6,16,16-4-Androsten-11β-ol-3,17-dione, D3-16,16,17-11-ketotestosterone. 250 µL of each sample, as well as 20 µL of the internal standard mixture or the appropriate quality controls were extracted by supported liquid extraction using methyl-tert-butyl-ether and then reconstituted with 40 % (v/v) methanol. Chromatography was carried out on a Waters HSS T3 1.8 µm, 2.1 x 50 mm column, with 2 mmol/L ammonium acetate, 0.1 % (v/v) formic acid in water as the aqueous mobile phase and acetonitrile as the organic mobile phase. Quantification by mass spectrometry was then conducted using a Waters XEVO TQ-XS system operated in positive ion mode. Standardized validation included a mean recovery of 92.5 - 109.8 for all analytes at low, medium and high concentration, an intra-assay coefficient of variation (26) of < 8.1 % with a bias between -6.6 and 4.2 and an inter-assay CV of < 11.4 % with a bias between -12.3 and 13.5 %. Lower limits of quantification (LLOQ) were defined as 12.5 pmol/L for 17OHP, 10 pmol/L for A4, 5 pmol/L for T, 45 pmol/L for 11OHA4 and 6 pmol/L for 11KT. The mean bias between direct sample analysis and overnight storage at 4 °C was 3.3 % for 17OHP, -2.6 % for A4, 1.2 % for T, 2.6 % for 11OHA4 and -1.8 % for 11KT. Carry-over at supra-physiological concentrations of each analyte was 0.07 % for 17OHP, 0.05 % for A4, 0.03 % for T, 0.007 % for 11OHA4 and 0.03 % for 11KT (20,25).

### Statistical analysis

For statistical analysis data was tested for normality using the Shapiro-Wilk test in conjunction with graphical display by histogram and Q-Q plot. Column statistics were calculated using GraphPad Prism (Mean, SEM, Quartiles). For longitudinal analysis hormone values were log-transformed before further processing due to their non-normal distribution. For reasons of clarity only untransformed values are reported. Due to the unbalanced design and missing datapoints, changes in salivary hormone values during the day were examined by a linear mixed-effects model for repeated measures with time as fixed and subject as random effect using an unstructured covariance structure (27). AUC was calculated with baseline Y = 0, positive peak direction and ignoring peaks that are less than 10% of the distance from minimum to maximum Y using GraphPad Prism. The percentage differences of mean hormone levels (Δ_mean_) were calculated by defining the fracture of evening hormone levels (time-point 5) / morning hormone levels (time-point 1) and calculating the percentage change. Difference of hormone levels between time-point 1 and 5 (Δ_hormone levels_) was identified by subtraction and presented as mean (SEM) in pg/ml. Differences in AUC were examined using the Mann-Whitney-U test. For correlation analysis Spearman’s correlation coefficient was computed for non-parametric data. The confidence interval was defined as 95% and a p-value of < 0.05 was considered statistically significant P ≤ 0.05 (≤ 0.05 (*), ≤ 0.01 (**), ≤ 0.001 (***)). Statistical analysis and graphical presentation were carried out using SPSS Statistics Software Version 26, GraphPad Prism 7.03 and Adobe Illustrator 2020.

## Results

### Characteristics of study participants

Median age was 29.5 years (IQR 23.8-37.3) in male (controls: 31.0 (25.0-42.0); p = 0.805) and 26.0 years (IQR 22.5-36.0) in female patients (controls: 26.0 (23.0-28.5; p = 0.911). Median BMI was 26.7 kg/m^2^ in men (IQR 24.7-29.5) and 27.6 kg/m^2^ in women with CAH (IQR (23.8-31.2), comparable to controls (Table 3).

**Table 3.**
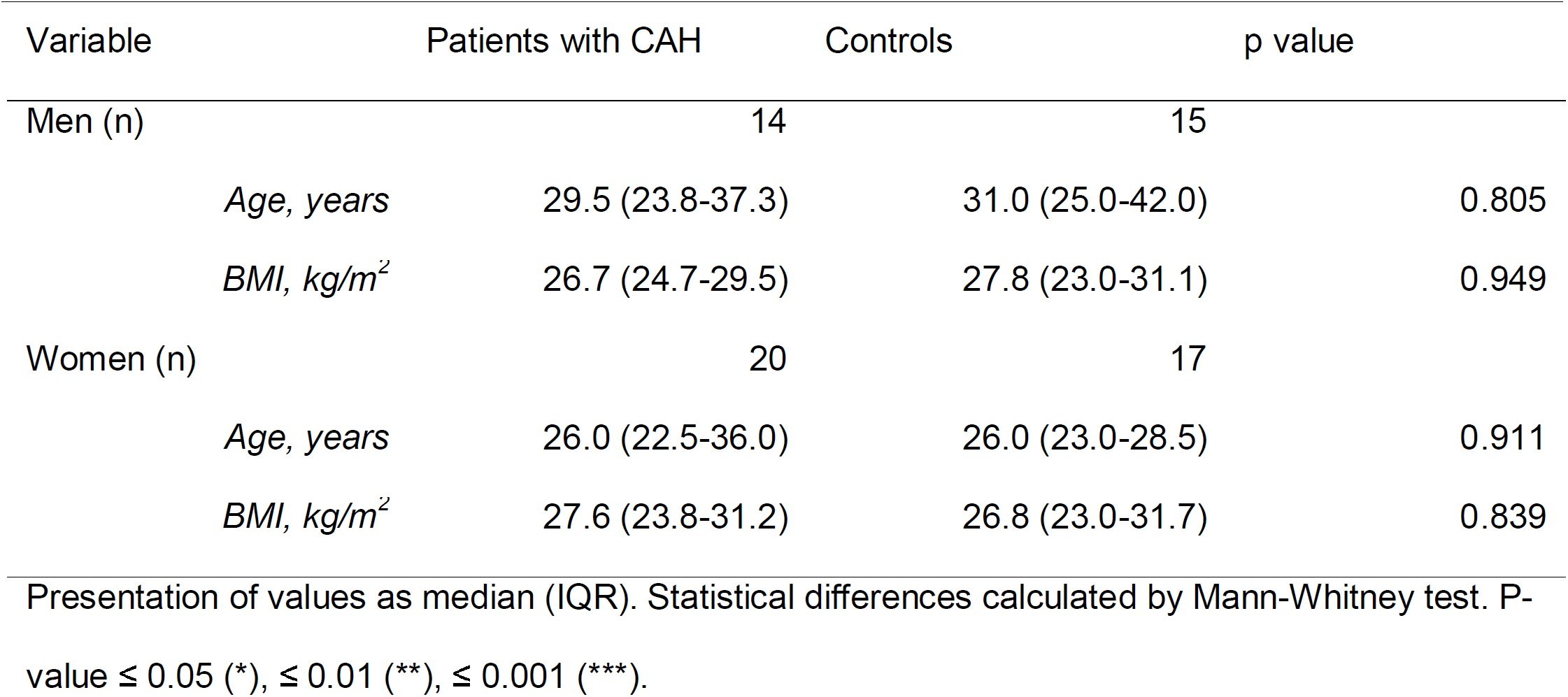
Baseline characteristics of patients with CAH and controls.

As depicted by Table 4, the majority of CAH patients received hydrocortisone (35.7 % of men and 55.0 % of women) or prednisolone (50.0 % of males and 40.0 % of females). Three patients (two men and one woman) received dexamethasone. The mean equivalent dose applied varied between 22.5 mg (13.10 mg/m^2^) in women and 30 mg (14.74 mg/m^2^) in men. All patients were on a circadian medication scheme, none of them substituted in a reverse-circadian manner (Table 4).

**Table 4.** Medication of patients with CAH.

|  | Men with CAH | Women with CAH |
| --- | --- | --- |
| Glucocorticoids |  |  |
| *Hydrocortisone* | 5 (35.7) | 11 (55.0) |
| *Prednisolone* | 7 (50.0) | 8 (40.0) |
| *Dexamethasone* | 2 (14.3) | 1 (5.0) |
| *Equivalent dosage, mg* | 30 (23.75 – 32.5) | 22.5 (20.0 – 29.0) |
| *Equivalent dosage, mg/m<sup>2</sup>* | 14.74 (12.30 – 18.04) | 13.10 (11.60 – 14.22) |
| *Circadian administration regimen* | 14 (100) | 20 (100) |
| Fludrocortisone | 9 (64.3) | 14 (70.0) |
| *Dosage, mg* | 0.1 (0.05-0.1375) | 0.075 (0.05-1.0) |

Presentation of values as median (IQR) or n (%).

According to biochemical and hormonal criteria, most patients were regarded as being well-controlled in terms of androgen excess with the exception of two male and two female patients. In the female CAH cohort, two patients were rather overtreated with suppressed androgen concentrations. Criteria for assessment of therapeutic control were A4 and T AUC, as well as A4/T AUC ratio within the range of the corresponding control group (6) (Table 2), while increased 17OHP AUC concentrations compared to controls were regarded as target for patients with classic 21OHD.

### Diurnal rhythm of 17OHP and (adrenal-specific) androgens

17OHP measurements showed a distinct diurnal rhythm with highest measurements in the early morning and declining values throughout the day in both CAH patients and healthy controls, as illustrated by Figure 2, Table 5 and 6 (male patients: F_1,11.841_ = 2.660; p = 0.085; Δ_mean_ = 75 %; male controls: F_1,13.630_ = 16.670; p < 0.001; Δ_mean_ = 42 %; female patients: F_1,16.842_ = 10.674; p < 000.1; Δ_mean_ = 82 %; female controls: F_1,13.040_ = 9.115; p = 0.001; Δ_mean_ = 51 %). However, the morning peaks in 17OHP concentrations clearly varied between patients and controls with a more than ten-fold increase in 17OHP in CAH patients. Correspondingly, for male patients with CAH, the AUC for 17OHP was significantly larger than for respective controls 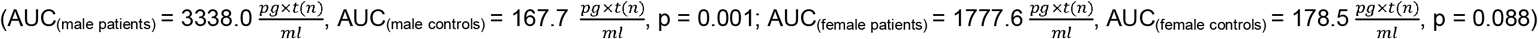.

**Table 5.**
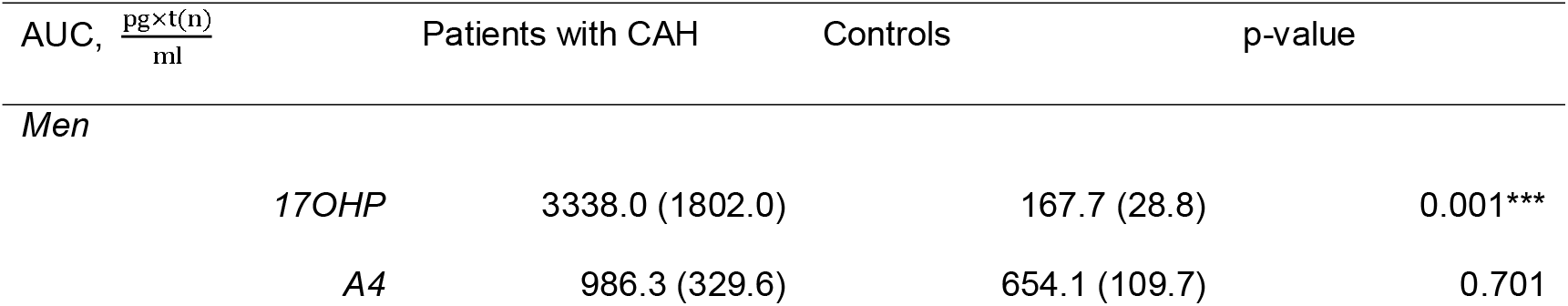
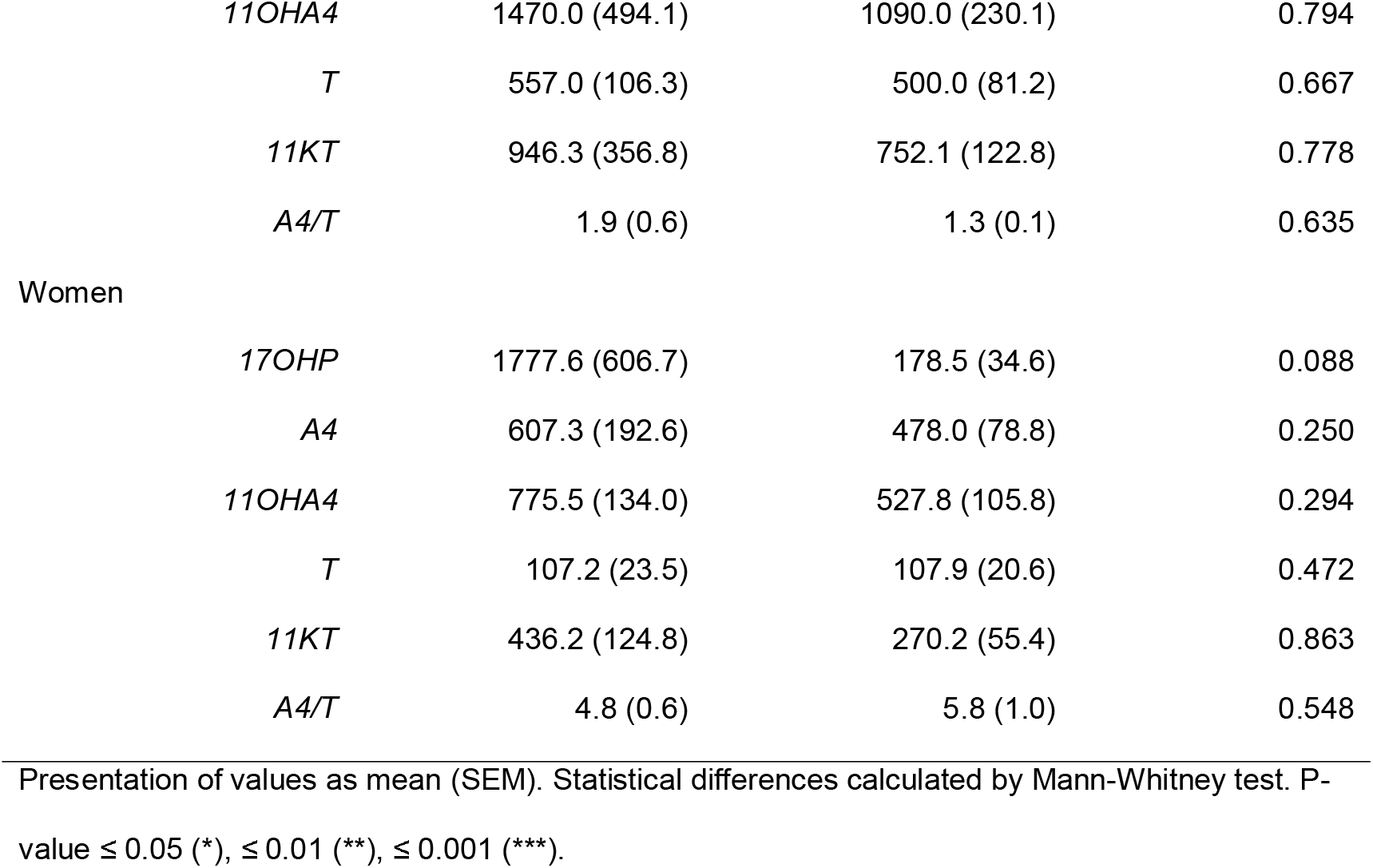
AUC of salivary hormone profiles of patients with CAH and controls.

| AUC, $\frac{\text{pg} \times \text{t(n)}}{\text{ml}}$ | Patients with CAH | Controls | p-value |
| --- | --- | --- | --- |
| *Men* |  |  |  |
| *17OHP* | 3338.0 (1802.0) | 167.7 (28.8) | 0.001*** |
| *A4* | 986.3 (329.6) | 654.1 (109.7) | 0.701 |

|  |  |  |  |  |
| --- | --- | --- | --- | --- |
|  | 11OHA4 | 1470.0 (494.1) | 1090.0 (230.1) | 0.794 |
|  | T | 557.0 (106.3) | 500.0 (81.2) | 0.667 |
|  | 11KT | 946.3 (356.8) | 752.1 (122.8) | 0.778 |
|  | A4/T | 1.9 (0.6) | 1.3 (0.1) | 0.635 |
| Women |  |  |  |  |
|  | 17OHP | 1777.6 (606.7) | 178.5 (34.6) | 0.088 |
|  | A4 | 607.3 (192.6) | 478.0 (78.8) | 0.250 |
|  | 11OHA4 | 775.5 (134.0) | 527.8 (105.8) | 0.294 |
|  | T | 107.2 (23.5) | 107.9 (20.6) | 0.472 |
|  | 11KT | 436.2 (124.8) | 270.2 (55.4) | 0.863 |
|  | A4/T | 4.8 (0.6) | 5.8 (1.0) | 0.548 |

Presentation of values as mean (SEM). Statistical differences calculated by Mann-Whitney test. P-value $\leq 0.05$ (\*), $\leq 0.01$ (\*\*), $\leq 0.001$ (\*\*\*).

**Table 6.**
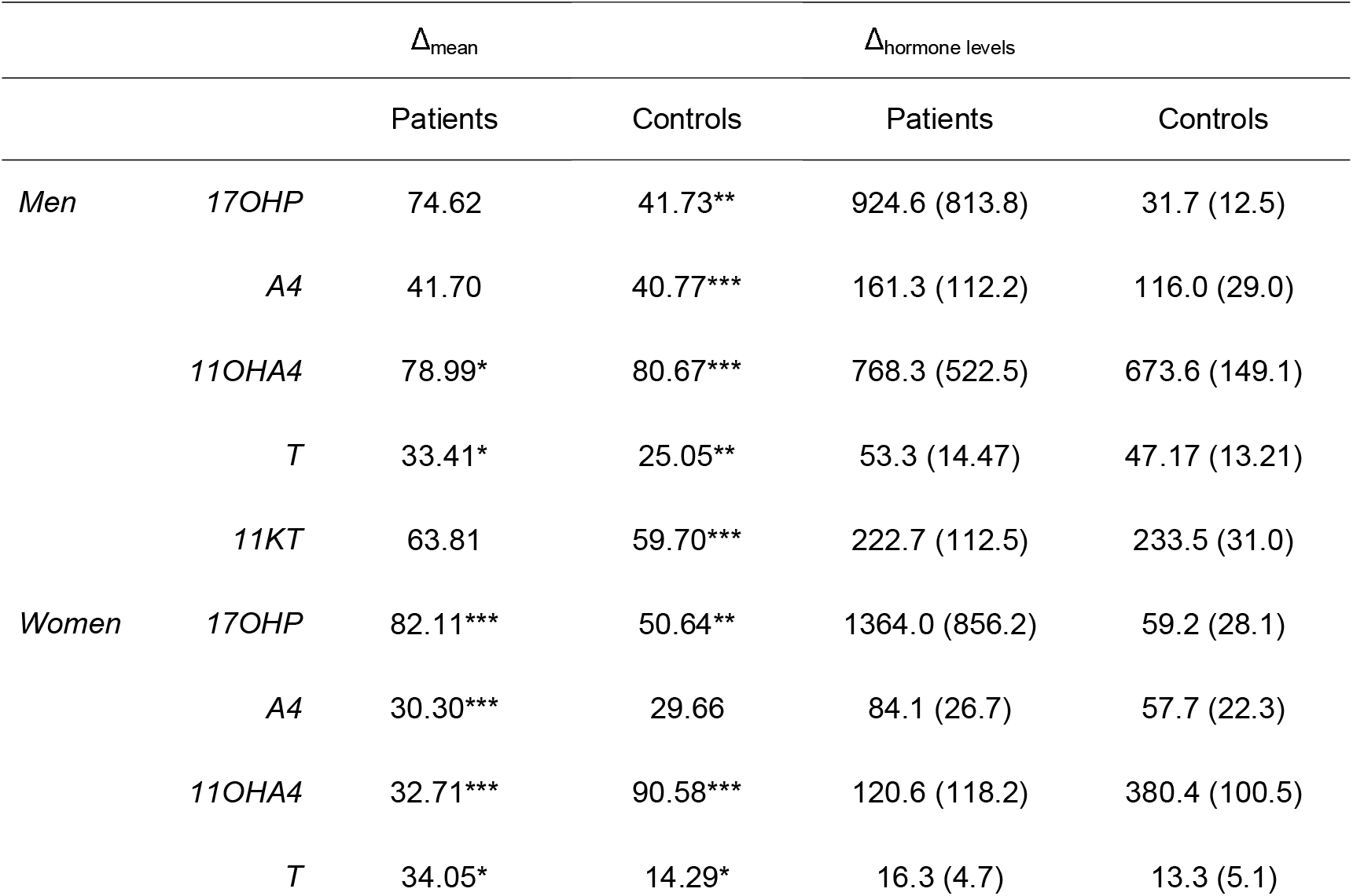
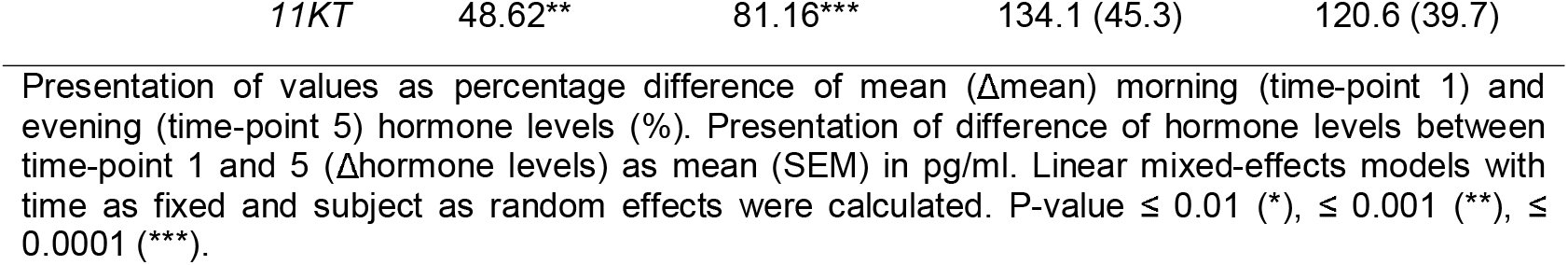
Mean percentage difference of morning and evening salivary hormone levels in patients with CAH and controls.

**Figure 2.**
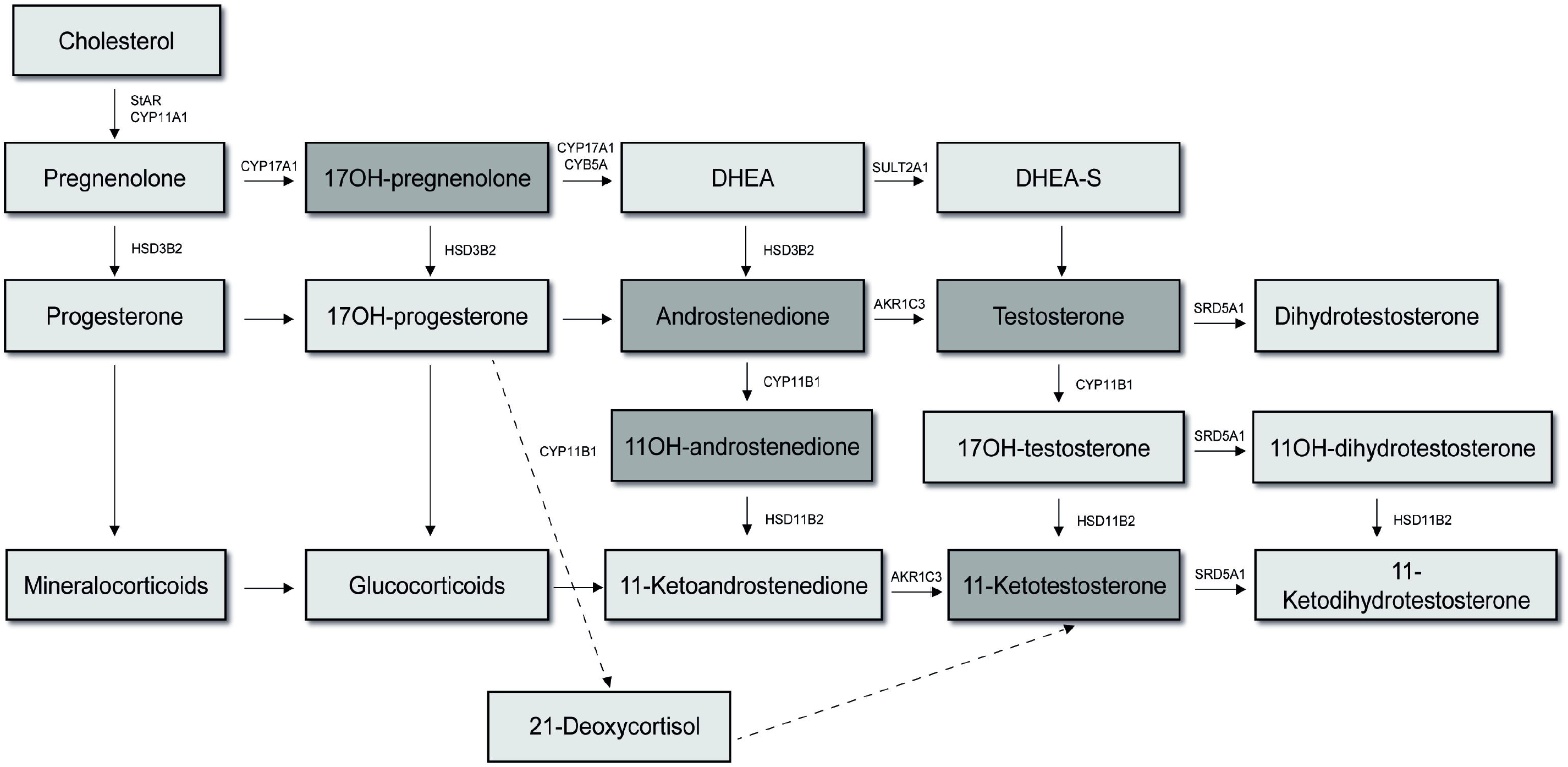
Diurnal variation of 17OHP, A4, 11OHA4, T and 11KT levels in saliva of patients with CAH and matched controls. Saliva profiles consist of measurements at up to five different timepoints throughout the day. Levels of 17OHP, A4, 11OHA4, T and 11KT were measured using LC-MS/MS. Hormone levels of patients with CAH are illustrated in red, data of respective controls in black colour. The mean of AUC of hormonal levels over the whole day are indicated per graph 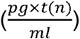. Please note that timepoints are not exactely the same in patients and controls.

A comparable diurnal rhythmicity could also be observed for A4, 11OHA4 and 11KT. Regarding A4, the mean changes in a diurnal pattern were equally pronounced in CAH patients compared to controls, while the inter-individual variance was higher in patients as expected (male patients: F_1,11.320_ = 2.298, p = 0.122, Δ_mean_ = 42 %; male controls: F_1,14.211_ = 21.849, p < 0.001, Δ_mean_ = 41 %; female patients: CAH women F_1,19.177_ = 10.797, Δ_mean_ = 30 %; female controls: F_1,4.478_ = 3.145, p = 0.133, Δ_mean_ = 30 %).

Overall, A4 secretion was not significantly different from controls 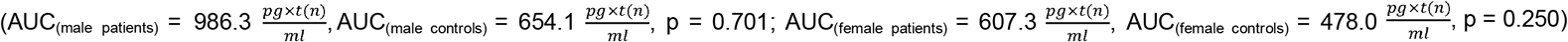.

11-OHA4 levels depicted a clear morning peak in both patients and control subgroups (male patients: F_1,12.051_ = 3.571, p = 0.038, Δ_mean_ = 79 %; male controls: F_1,13.409_ = 55.993, p < 0.001, Δ_mean_ = 81%; female patients: F_1,15.920_ = 12.912, p < 0.001, Δ_mean_ = 33 %; female controls: F_1. 21.085_ = 25.971, p < 0.001, Δ_mean_ = 91 %) with a nighttime increase only in female patients with CAH. Difference in the AUC appeared larger in the male cohort compared to female subjects but failed to reach clinical significance 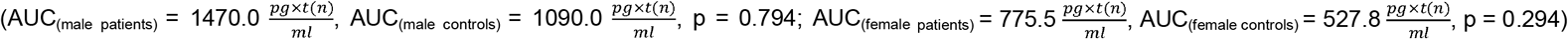.

Similarly, 11KT measurements also followed a diurnal rhythm with concentrations constantly declining over the course of the day following an early morning peak (male patients: F_1,7.872_ = 3.686. p = 0.056; Δ_mean_ = 64 %; male controls: F_1,13.702_ = 25.139; p < 0.001; Δ_mean_ = 60 %; female patients: F_1,15.942_ = 8.349; p = 0.001; Δ_mean_ = 49 %; female controls: F_1,13.225_ = 35.107; p < 0.001; Δ_mean_ = 81 %). An early rise towards the night was only present in female patients with CAH, although there was no difference in intake of evening GC dose between men and women. There was no significant difference in the corresponding AUC between groups of either sex 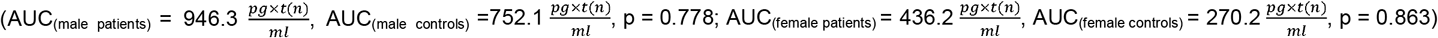.

The smallest degree of diurnal variation was seen in T measurements (male patients: F_1,12.619_ = 5.935, p = 0.006, Δ_mean_ = 33 %; male controls: F_1,12.888_ = 9.763, p = 0.001, Δ_mean_ = 25 %; female patients: F_1,12.100_ = 5.832, p = 0.007, Δ_mean_ = 34 %; female controls: F_1, 8.585_ = 4.596, p = 0.029, Δ_mean_ = 14 %). Testosterone AUC was higher in males versus females, no statistical difference in AUC was observed between male and female patients and controls (AUC_(male patients)_ = 557.0, AUC_(male controls)_ = 500.0, p = 0.667; AUC_(female patients)_ = 107.2, AUC_(female controls)_ = 107.9, p = 0.472).

### Correlation of (adrenal-specific) androgens with 17-OHP in healthy controls

Control subjects showed significant correlations of 17OHP- and T-AUC (r(p)_male_ = 0.622*; r(p)_female_ = 0.648*) and male controls also a significant correlation of the AUC of 17OHP and A4 (r(p)_male_ = 0.855***; r(p)_female_ = 0.429) as depicted by Table 7 and 8. In contrast, 11KT (r(p)_male_ = 0.345; r(p)_female_ = 0.532) and 11OH4 (r(p)_male_ = 0.429; r(p)_female_ = -0.192) did not significantly correlate with 17OHP concentrations in saliva.

**Table 7.**
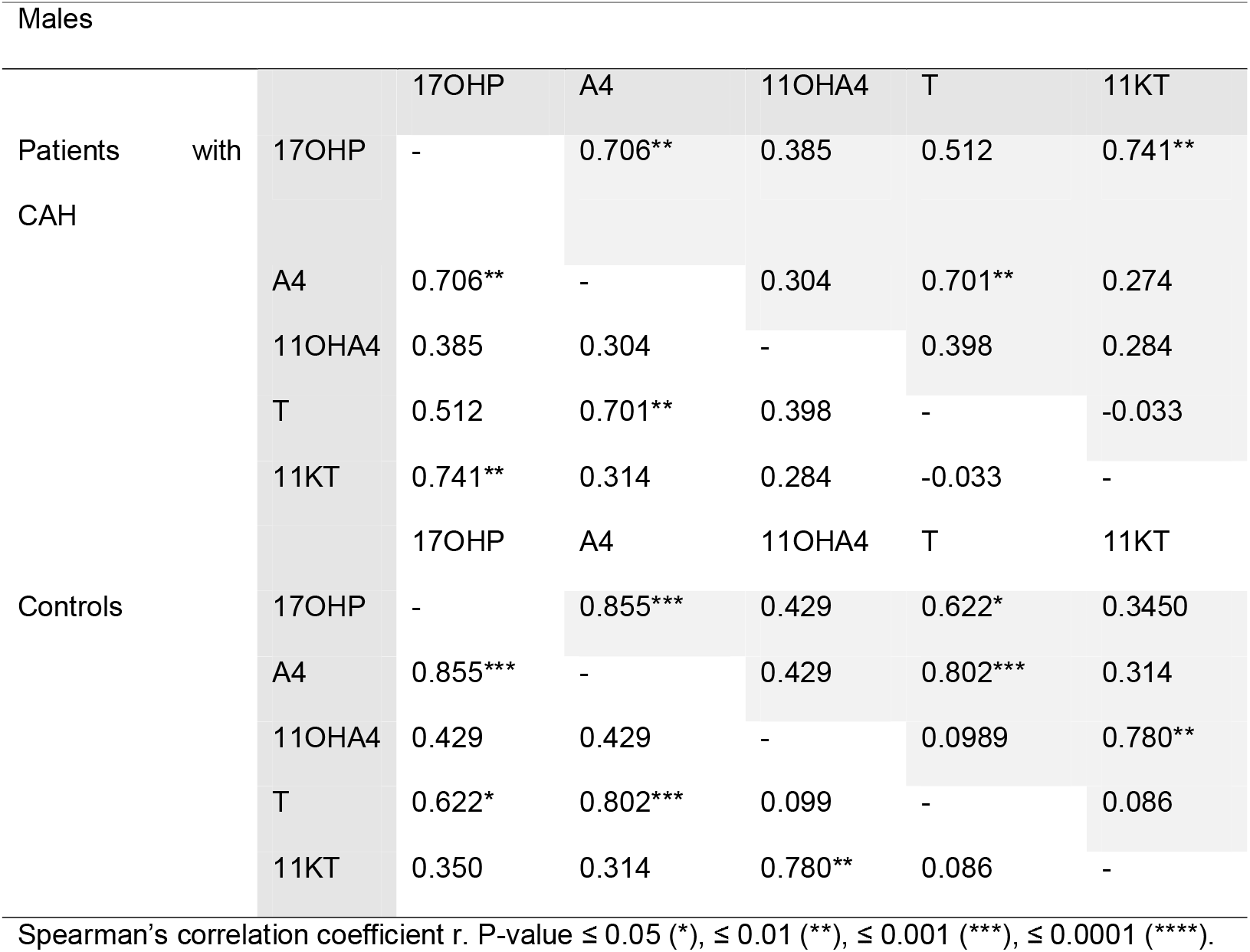
Correlation of AUC in male patients with CAH and controls.

**Table 8.**
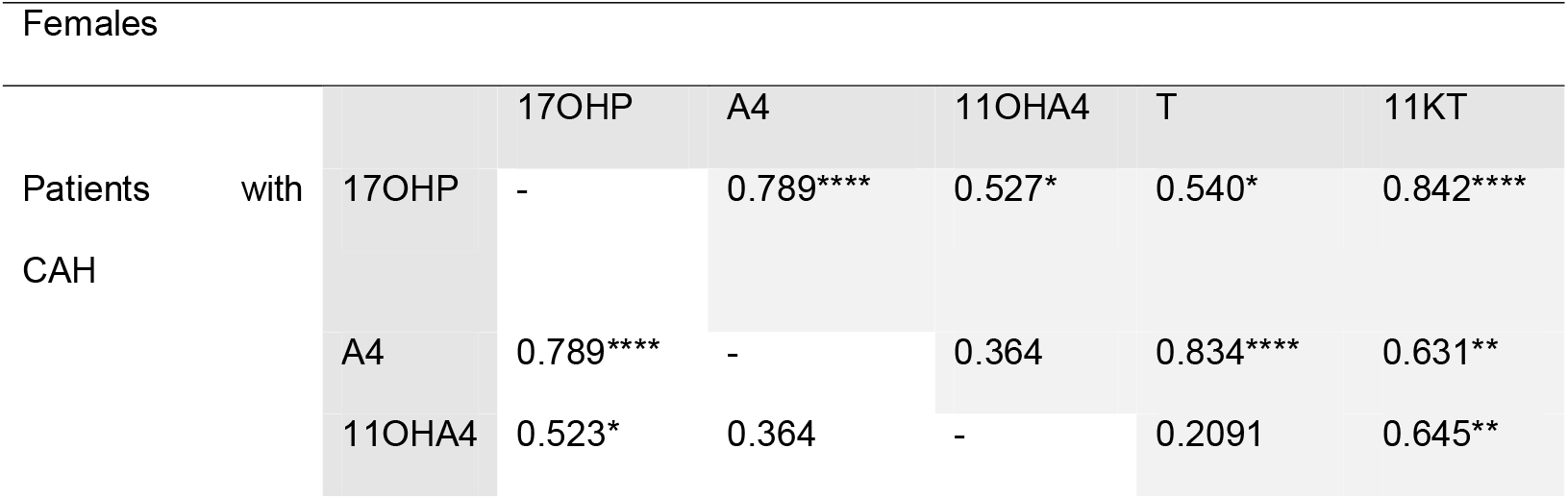
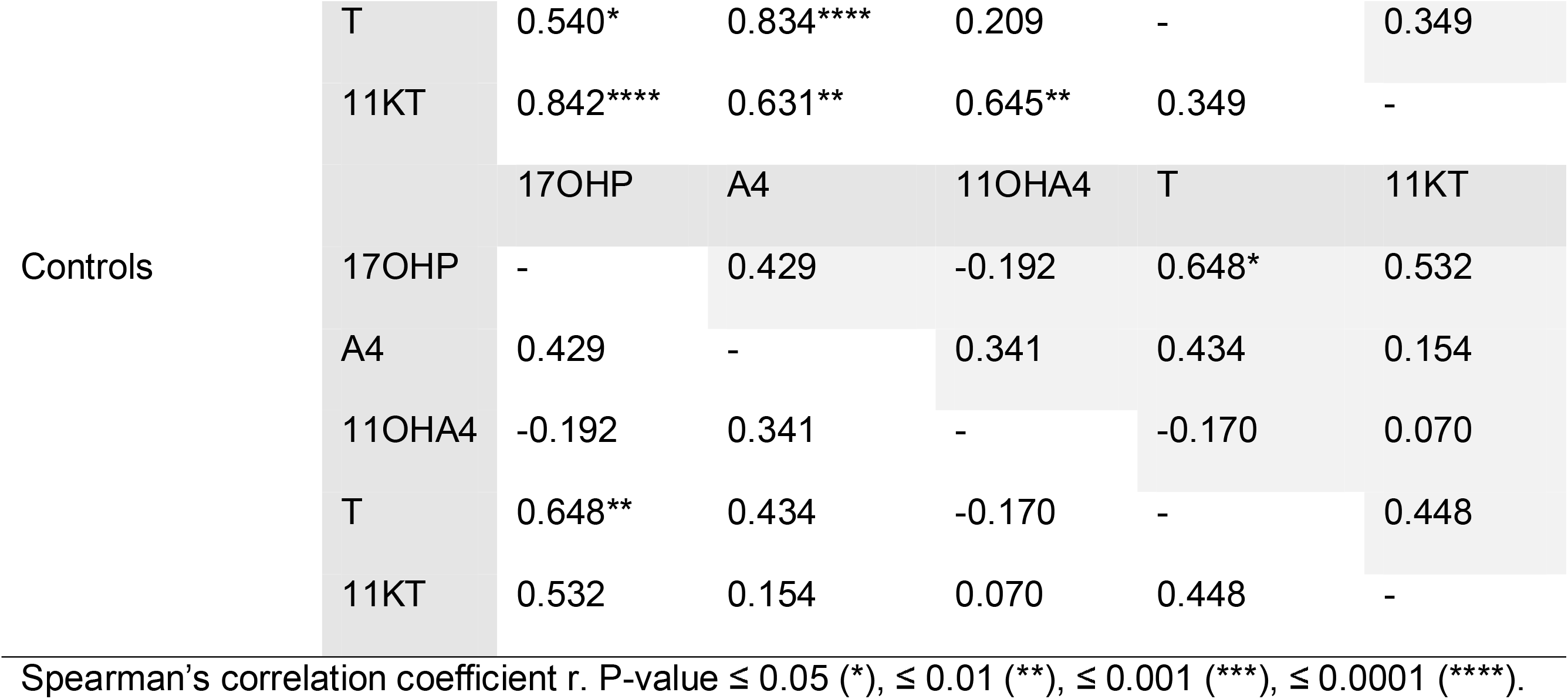
Correlation of AUC in female patients with CAH and controls.

In patients, the correlation between 17OHP measurements in saliva samples and T was weaker compared to controls (r(p)_male_ = 0.512; r(p)_female_ = 0.540*). 17OHP and A4 measurements in CAH patients presented with a very strong correlation throughout the whole day (r(p)_male_ = 0.706**; r(p)_female_ = 0.789***). Regarding the measured 11oxC19 androgens, a strong correlation could be observed between 11KT and 17OHP in both male and female patients (r(p)_male_ = 0.741**; r(p)_female_ = 0.842***), as well as a significant correlation for 11OHA4 and 17OHP measurements in female patients (r(p)_male_ = 0.385^n.s.^; r(p)_female_ = 0.527*).

When correlating T and 11KT measurements in saliva samples of male patients with 21OHD, a negative trend, but no significant correlation was observed (r(p)_male_ = -0.033).

## Discussion

This is the first study to show that there is a clear circadian pattern for the investigated 11oxC19 androgens in the general population that is principally preserved or accentuated in patients with 21OHD. Our analysis therefore indicates that therapeutic monitoring of patients with 21OHD by measurement of 11oxC19 steroids in saliva might be an efficient method, if using salivary profiles at multiple timepoints throughout the day.

A diurnal profile with decreasing values from morning to evening has already been described for 17OHP, A4 and T (8,23,28). This physiological circadian rhythm in adrenal steroid hormone production is mainly driven by the rhythmic release of ACTH, in response to corticotropin releasing hormone (CRH) and vasopressin secretion from the paraventricular nucleus (PVN), which is under the control of the master clock in the suprachiasmatic nucleus (SCN) (29). In addition, there is also a peripheral adrenal clock that is tightly regulated by adrenal gland-specific time-controlled genes, such as steroidogenic acute regulatory protein (StAR) (30). As StAR is involved in the rate limiting step of steroid production, this results in a robust circadian rhythm of adrenal steroid synthesis (31). Here, we have shown that this diurnal variation also applies to 11oxC19 steroid production following the pattern of their precursor 17OHP (32).

Thus, 11oxC19 concentrations need to be interpreted with consideration of their diurnal variance and sampling time. Furthermore, GC pharmacokinetics as well as circadian or reverse circadian replacement regimen will impact on 11oxC19 steroid concentration. Due to low sample size we could not analyze the effects of different modes of GC replacement in our study. It will be highly interesting to explore the effects of immediate-versus modified-release hydrocortisone, intermediate and long-term synthetic GC on salivary steroid day profiles in future studies.

It is yet unclear if 11oxC19 steroids have an advantage over established markers regarding treatment outcomes. So far, there is no interventional study available exploring a potential difference in regard to clinical outcomes by targeting 11oxC19 steroid levels in comparison to other markers such as A4 or the A4/T-ratio (6,33). Given the close relationship of these steroids also raises the question if these steroids can be independently targeted by treatment interventions (12,18). Conversely, there are several studies suggesting that 11oxC19 steroids provide a major amount of additional information in 21OHD treatment monitoring and may be more suitable in adjusting GC treatment than currently used biomarkers.

The current treatment goal has been defined by normalized androgens while elevated 17OHP has been suggested as treatment target. It is generally accepted that while poorly controlled patients with 21OHD present with hugely elevated 17OHP levels, aiming for the reference range of healthy controls results in overtreatment (6). It has been shown that while A4 and T are normalized in clinically well-controlled patients with 21OHD, 17OHP concentrations are still elevated (34,35). In our study,11OHA4 and 11KT secretion did not significantly differ from our control population throughout the day despite significant inter-individual variations and excessively elevated 17OHP concentrations in patients (12,18). This underlines that the utility of 17OHP as a treatment marker is restricted to detect overtreatment.

Comparable A4, T and 11oxC19 androgens in patients and controls over the course of the day in our study confirm that according to our initial assessment based on a single measurement of serum steroids, overall disease control in our cohort was considered to be very good. This notion is further supported by the fact that there was no significant difference in the AUC of A4 and T nor the AUC A4/T ratio between patients and controls in our study (Table 5). Accordingly, it has been shown before that 11oxC19 androgen measurements in patients with 21OHD show clear overlap with those of healthy controls (20). Finally, a significant correlation of salivary 17OHP and 11oxC19 androgen measurements was observed in patients but not in healthy controls (Table 7), indicating the limited relevance of the 11oxC19 pathway in subjects with a functioning enzymatic pathway of steroidogenesis (5).

A potential major advantage of 11oxC19 steroids over classic androgens in 21OHD might be that they have proven adrenal origin and therefore might be better linked with clinical outcome markers (17). Consequently, in patients with 21OHD, 11oxC19 concentrations were found to be associated with increased adrenal volume, menstrual disturbances in women and the presence of testicular adrenal rest tumor in males (17,18). A particular challenge in disease monitoring is that the currently used classic biomarkers A4 and T are derived from both the adrenals and the gonads and their source cannot be distinguished by laboratory methods. To help distinguish T from adrenal or testicular origin in male patients with 21OHD it has been suggested to use the serum A4/T ratio (36). This is based on the observation that A4 is elevated when androgens are predominantly of adrenal origin and result in suppression of gonadotrophins and an elevated A4/T ratio. This is supported by the observation of Turcu et al. who described a clear significant inverse correlation between T and 11KT measurements in serum. It was hypothesized that due to the strong androgenic activity of 11KT, excessive secretion results in suppression of the hypothalamic-pituitary-gonadal (HPG) axis in males with 21OHD (18). This inverse correlation could not be confirmed in our study, albeit we observed a trend towards the inverse correlation of T and 11KT. These discrepant results could be explained by the fact that, at the timepoint of saliva sampling, most of the analyzed patients were under optimal hormonal control, limiting the variance in T and 11KT concentrations during the course of the day. Analysis of 11oxC19 androgens would allow direct measurement of adrenal-derived androgens and salivary day profiling will enable an integrated and coherent picture of adrenal steroid synthesis throughout the day. In particular, salivary day profiling in 21OHD patients will help to overcome the challenge of precisely timed blood sampling. Instead, it allows serial and more frequent day profiling in a non-invasive and patient-friendly manner. In addition, it allows monitoring in a telemedicine setting providing access to specialized tertiary care in this rare disease even in remote areas and in situations with travel restrictions as the current pandemic situation.

Due to very good disease control in almost all patients of the study and hardly any cases with undertreatment, limitations of this study were defined by a high percentage of measurements within the LLOQ. Clinical use of ultra-performance convergence chromatography tandem mass spectrometry (UPC^2^-MS/MS) for example could offer superior sensitivity resulting in even lower LOQs (37). In addition, the timepoints of sample collection in patients and controls were not identical in this study, leading to limited comparability of individual timepoints of data collection. However, our main goal in this study was to show diurnal variations within and not between groups. Finally, given the sample size, impact of different GC medications, immediate-versus modified-release GC, intermediate or long-acting synthetic GC, could not be evaluated in this study. Fluctuations in salivary androgens in relation and response to different GC replacement regimen therefore could not be analyzed in this study and need to be explored in the future.

To conclude, this study is the first to describe the diurnal rhythm of 11oxC19 androgen concentrations in salivary profiles in both healthy controls and patients with 21OHD. Our findings highlight that diurnal fluctuations of 11oxC19 need to be considered for the assessment of biochemical disease control in 21OHD. Ultimately, comprehensive salivary day steroid profiling will improve patient management and allow more frequent hormone measurements leading to more timely and accurate GC treatment adjustments.

## Data Availability

Data can be provided by the corresponding author on reasonable request

## Abbreviations

11β-HSD2: 11β-hydroxysteroid dehydrogenase type 2
11oxC19: 11-oxygenated 19-carbon
11OHA4: 11β-hydroxyandrostenedione
11KA4: 11-ketoandrostenedione
11KDHT: 11-ketodihydrotestosterone
11KT: 11-ketotestosterone
11OHT: 11-hydroxytestosterone
21DF: 21-deoxycortisol
21OHD: 21-hydroxylase deficiency
A4: androstenedione
ACTH: adrenocorticotropic hormone
AUC: area under the curve
CAH: congenital adrenal hyperplasia
CRH: corticotropin releasing hormone
CYP11B1: 11β-hydroxylase
DHT: dihydrotestosterone
GC: glucocorticoid
HPA: hypothalamic-pituitary-axis
HPG: hypothalamic-pituitary-gonadal
LLOQ: lowest limits of quantification
LP-MS/MS: liquid chromatography-mass spectrometry
PVN: paraventricular nucleus
StAR: steroidogenic acute regulatory protein
SCN: suprachiasmatic nucleus
T: testosterone
TART: testicular adrenal rest tumors
UPC^2^-MS/MS: ultra-performance convergence chromatography tandem mass spectrometry

## Acknowledgments

We thank all participants of this study.

## Author Contributions

N.R. designed the study, H.N and M.K.A. contributed to the study concept. C.L., H.S. and I.D. provided data. H.N. and M.K.A. conducted the statistical analysis. M.B. J.A. and J.H. performed the laboratory analyses. H.N., M.K.A. and N.R. drafted the manuscript. All authors provided intellectual input and read, revised and approved the final version of the manuscript.

## Data Availability

The datasets generated during and/or analyzed during the current study are not publicly available but are available from the corresponding author on reasonable request.

## References

1. Ali SR, Bryce J, Haghpanahan H, Lewsey JD, Tan LE, Atapattu N, Birkebaek NH, Blankenstein O, Neumann U, Balsamo A, Ortolano R, Bonfig W, Claahsen-van der Grinten HL, Cools M, Costa EC, Darendeliler F, Poyrazoglu S, Elsedfy H, Finken MJJ, Fluck CE, Gevers E, Korbonits M, Guaragna-Filho G, Guran T, Guven A, Hannema SE, Higham C, Hughes IA, Tadokoro-Cuccaro R, Thankamony A, Iotova V, Krone NP, Krone R, Lichiardopol C, Luczay A, Mendonca BB, Bachega T, Miranda MC, Milenkovic T, Mohnike K, Nordenstrom A, Einaudi S, van der Kamp H, Vieites A, de Vries L, Ross RJM, Ahmed SF. Real World Estimates Of Adrenal Insufficiency Related Adverse Events In Children With Congenital Adrenal Hyperplasia. J Clin Endocrinol Metab. 2020.

2. Therrell BL. Newborn screening for congenital adrenal hyperplasia. Endocrinol Metab Clin North Am. 2001;30(1):15–30.

3. Kamrath C, Hochberg Z, Hartmann MF, Remer T, Wudy SA. Increased activation of the alternative “backdoor” pathway in patients with 21-hydroxylase deficiency: evidence from urinary steroid hormone analysis. J Clin Endocrinol Metab. 2012;97(3):E367–375.

4. Pignatelli D, Pereira SS, Pasquali R. Androgens in Congenital Adrenal Hyperplasia. Front Horm Res. 2019;53:65–76.

5. Turcu AF, Auchus RJ. Adrenal steroidogenesis and congenital adrenal hyperplasia. Endocrinol Metab Clin North Am. 2015;44(2):275–296.

6. Speiser PW, Arlt W, Auchus RJ, Baskin LS, Conway GS, Merke DP, Meyer-Bahlburg HFL, Miller WL, Murad MH, Oberfield SE, White PC. Congenital Adrenal Hyperplasia Due to Steroid 21-Hydroxylase Deficiency: An Endocrine Society Clinical Practice Guideline. J Clin Endocrinol Metab. 2018;103(11):4043–4088.

7. Shibayama Y, Higashi T, Shimada K, Kashimada K, Onishi T, Ono M, Miyai K, Mizutani S. Liquid chromatography-tandem mass spectrometric method for determination of salivary 17alpha-hydroxyprogesterone: a noninvasive tool for evaluating efficacy of hormone replacement therapy in congenital adrenal hyperplasia. J Chromatogr B Analyt Technol Biomed Life Sci. 2008;867(1):49–56.

8. Juniarto AZ, Goossens K, Setyawati BA, Drop SL, de Jong FH, Faradz SM. Correlation between androstenedione and 17-hydroxyprogesterone levels in the saliva and plasma of patients with congenital adrenal hyperplasia. Singapore Med J. 2011;52(11):810–813.

9. Bloem LM, Storbeck KH, Swart P, du Toit T, Schloms L, Swart AC. Advances in the analytical methodologies: Profiling steroids in familiar pathways-challenging dogmas. J Steroid Biochem Mol Biol. 2015;153:80–92.

10. de Groot MJ, Pijnenburg-Kleizen KJ, Thomas CM, Sweep FC, Stikkelbroeck NM, Otten BJ, Claahsen-van der Grinten HL. Salivary morning androstenedione and 17alpha-OH progesterone levels in childhood and puberty in patients with classic congenital adrenal hyperplasia. Clin Chem Lab Med. 2015;53(3):461–468.

11. Storbeck K-H, Schiffer L, Baranowski ES, Chortis V, Prete A, Barnard L, Gilligan LC, Taylor AE, Idkowiak J, Arlt W. Steroid metabolome analysis in disorders of adrenal steroid biosynthesis and metabolism. Endocrine Reviews. 2019;40(6):1605–1625.

12. Kamrath C, Wettstaedt L, Boettcher C, Hartmann MF, Wudy SA. Androgen excess is due to elevated 11-oxygenated androgens in treated children with congenital adrenal hyperplasia. J Steroid Biochem Mol Biol. 2018;178:221–228.

13. Jha S, El-Maouche D, Marko J, Mallappa A, Veeraraghavan P, Merke DP. Individualizing Management of Infertility in Classic Congenital Adrenal Hyperplasia and Testicular Adrenal Rest Tumors. J Endocr Soc. 2019;3(12):2290–2294.

14. Auer MK, Krumbholz A, Bidlingmaier M, Thieme D, Reisch N. Steroid 17-Hydroxyprogesterone in Hair Is a Potential Long-Term Biomarker of Androgen Control in Congenital Adrenal Hyperplasia due to 21-Hydroxylase Deficiency. Neuroendocrinology. 2020;110(11-12):938–949.

15. Dauber A, Kellogg M, Majzoub JA. Monitoring of therapy in congenital adrenal hyperplasia. Clin Chem. 2010;56(8):1245–1251.

16. Dahl SR, Nermoen I, Bronstad I, Husebye ES, Lovas K, Thorsby PM. Assay of steroids by liquid chromatography-tandem mass spectrometry in monitoring 21-hydroxylase deficiency. Endocr Connect. 2018;7(12):1542–1550.

17. Turcu AF, Mallappa A, Elman MS, Avila NA, Marko J, Rao H, Tsodikov A, Auchus RJ, Merke DP. 11-Oxygenated Androgens Are Biomarkers of Adrenal Volume and Testicular Adrenal Rest Tumors in 21-Hydroxylase Deficiency. J Clin Endocrinol Metab. 2017;102(8):2701–2710.

18. Turcu AF, Nanba AT, Chomic R, Upadhyay SK, Giordano TJ, Shields JJ, Merke DP, Rainey WE, Auchus RJ. Adrenal-derived 11-oxygenated 19-carbon steroids are the dominant androgens in classic 21-hydroxylase deficiency. Eur J Endocrinol. 2016;174(5):601–609.

19. Pretorius E, Arlt W, Storbeck KH. A new dawn for androgens: Novel lessons from 11-oxygenated C19 steroids. Mol Cell Endocrinol. 2017;441:76–85.

20. Bacila I, Adaway J, Hawley J, Mahdi S, Krone R, Patel L, Alvi S, Randell T, Gevers E, Dattani M, Cheetham T, Kyriakou A, Schiffer L, Ryan F, Crowne E, Davies JH, Ahmed SF, Keevil B, Krone N. Measurement of Salivary Adrenal-Specific Androgens as Biomarkers of Therapy Control in 21-Hydroxylase Deficiency. J Clin Endocrinol Metab. 2019;104(12):6417–6429.

21. Ruggiero C, Lalli E. Impact of ACTH Signaling on Transcriptional Regulation of Steroidogenic Genes. Front Endocrinol (Lausanne). 2016;7:24–24.

22. HPA Axis-Rhythms. Comprehensive Physiology:1273–1298.

23. Mezzullo M, Fazzini A, Gambineri A, Di Dalmazi G, Mazza R, Pelusi C, Vicennati V, Pasquali R, Pagotto U, Fanelli F. Parallel diurnal fluctuation of testosterone, androstenedione, dehydroepiandrosterone and 17OHprogesterone as assessed in serum and saliva: validation of a novel liquid chromatography-tandem mass spectrometry method for salivary steroid profiling. Clin Chem Lab Med. 2017;55(9):1315–1323.

24. Vogel F, Braun LT, Rubinstein G, Zopp S, Künzel H, Strasding F, Albani A, Riester A, Schmidmaier R, Bidlingmaier M, Quinkler M, Deutschbein T, Beuschlein F, Reincke M. Persisting Muscle Dysfunction in Cushing’s Syndrome Despite Biochemical Remission. The Journal of clinical endocrinology and metabolism. 2020;105(12):dgaa625.

25. Schiffer L, Adaway JE, Arlt W, Keevil BG. A liquid chromatography-tandem mass spectrometry assay for the profiling of classical and 11-oxygenated androgens in saliva. Ann Clin Biochem. 2019;56(5):564–573.

26. Deterding K SC, Schott E, Welzel TM, Gerken G, Klinker H, Spengler U, Wiegand J, Schulze Zur Wiesch J, Pathil A, Cornberg M, Umgelter A, Zöllner C, Zeuzem S, Papkalla A, Weber K, Hardtke S, von der Leyen H, Koch A, von Witzendorff D, Manns MP, Wedemeyer H; HepNet Acute HCV IV Study Group. Ledipasvir plus sofosbuvir fixed-dose combination for 6 weeks in patients with acute hepatitis C virus genotype 1 monoinfection (HepNet Acute HCV IV): an open-label, single-arm, phase 2 study. Lancet Infectious Diseases. 2017;17(2):215–222.

27. Krueger C, Tian L. A Comparison of the General Linear Mixed Model and Repeated Measures ANOVA Using a Dataset with Multiple Missing Data Points. Biological Research For Nursing. 2004;6(2):151–157.

28. Debono M, Mallappa A, Gounden V, Nella AA, Harrison RF, Crutchfield CA, Backlund PS, Soldin SJ, Ross RJ, Merke DP. Hormonal circadian rhythms in patients with congenital adrenal hyperplasia: identifying optimal monitoring times and novel disease biomarkers. European journal of endocrinology. 2015;173(6):727–737.

29. Oster H, Challet E, Ott V, Arvat E, de Kloet ER, Dijk DJ, Lightman S, Vgontzas A, Van Cauter E. The Functional and Clinical Significance of the 24-Hour Rhythm of Circulating Glucocorticoids. Endocr Rev. 2017;38(1):3–45.

30. Son GH, Chung S, Choe HK, Kim H-D, Baik S-M, Lee H, Lee H-W, Choi S, Sun W, Kim H, Cho S, Lee KH, Kim K. Adrenal peripheral clock controls the autonomous circadian rhythm of glucocorticoid by causing rhythmic steroid production. Proc Natl Acad Sci U S A. 2008;105(52):20970–20975.

31. Chung S, Son GH, Kim K. Circadian rhythm of adrenal glucocorticoid: its regulation and clinical implications. Biochim Biophys Acta. 2011;1812(5):581–591.

32. Turcu AF, Rege J, Auchus RJ, Rainey WE. 11-Oxygenated androgens in health and disease. Nat Rev Endocrinol. 2020;16(5):284–296.

33. Auchus RJ, Arlt W. Approach to the patient: the adult with congenital adrenal hyperplasia. J Clin Endocrinol Metab. 2013;98(7):2645–2655.

34. Lee PA, Urban MD, Gutai JP, Migeon CJ. Plasma Progesterone, 17-Hydroxyprogesterone, Androstenedione and Testosterone in Prepubertal, Pubertal and Adult Subjects with Congenital Virilizing Adrenal Hyperplasia as Indicators of Adrenal Suppression. Hormone Research in Paediatrics. 1980;13(6):347–357.

35. Bode HH, Rivkees SA, Cowley DM, Pardy K, Johnson S. Home monitoring of 17 hydroxyprogesterone levels in congenital adrenal hyperplasia with filter paper blood samples. The Journal of pediatrics. 1999;134(2):185–189.

36. Auchus RJ. Management considerations for the adult with congenital adrenal hyperplasia. Mol Cell Endocrinol. 2015;408:190–197.

37. Quanson JL, Stander MA, Pretorius E, Jenkinson C, Taylor AE, Storbeck KH. High-throughput analysis of 19 endogenous androgenic steroids by ultra-performance convergence chromatography tandem mass spectrometry. J Chromatogr B Analyt Technol Biomed Life Sci. 2016;1031:131–138.

